# Exercise Training Improves Cognitive Function and Neurovascular Control in Patients With Heart Failure and Atrial Fibrillation: A Randomized Clinical Trial

**DOI:** 10.1101/2024.04.29.24306591

**Authors:** Guilherme Veiga Guimarães, Paulo Roberto Chizzola, Veridiana Moraes D’Avila, Paulo Roberto Santo-Silva, Raphaela Villar Grohes Miranda, Leandro Silva Alves, Edimar Alcides Bocchi

## Abstract

**Background:** Atrial fibrillation (AF) may be linked to cognitive impairment in heart failure (HF) patients. Regular exercise training has numerous benefits, including improving adults’ overall health and cognitive function. However, the effects of exercise training on cognitive function in heart failure patients with reduced ejection fraction and permanent atrial fibrillation (HFAF) are not well understood.

**Objective:** To test the hypothesis that exercise training improves cognitive function and neurovascular control in patients with HFAF.

**Methods:** This randomized clinical trial included patients with HFAF, LVEF ≤40%, and resting HR ≤80 bpm. Montreal Cognitive Assessment score (MoCA), muscle sympathetic nerve activity (MSNA), and forearm blood flow (FBF) assessment were performed before and after the 12-week protocol period.

**Results:** Twenty-six patients were randomized to exercise training (HFAF-trained, n=13) or no training (HFAF-untrained, n=13). At baseline, no differences between groups were found. Exercise significantly improved the MoCA score from 14.85 to 22.62 (*P*<0.0001), as well as those measuring processing visuospatial, memory, and attention function from 2.23 to 3.31 (*P*=0.02); from 1.92 to 2.69 (*P*=0.01); and from 2.62 to 3.85 (*P*=0.002), respectively. HFAF-training significantly decreased MSNA from 49.6±9.5 to 32.0±7.2 burst/min (*P*<0.0001) and forearm vascular resistance from 48.8±13.1 to 28.1±7 units (*P*<0.0001). Concomitantly, FBF increased from 1.9±0.5 to 3.0±0.7 mL/min/100mL (*P*<0.0001). The untrained group increased MSNA from 50.9±10.6 to 57.6±9.2 burst/min (*P*=0.004). No significant changes were found in the other variables in untrained patients.

**Conclusion:** Exercise training improves cognitive function and neurovascular control in patients with heart failure with reduced ejection fraction and permanent atrial fibrillation.

## Introduction

Heart failure (HF) is a systemic disease not limited to the heart but affects other organ systems, including the brain.^1,2^ Atrial fibrillation (AF) is the most common sustained arrhythmia and is a well-known and common complication in patients with HF, where it may be present in up to 25% to 40% of patients.^1,2^ Both HF and AF patients have some degree of cognitive impairment, which is the internal mental process that underlies how people perceive, remember, speak, think, make decisions, and solve problems.^3^ Cognitive decline interferes with patient self-care and treatment adherence, which leads to higher rates of hospitalization and increased mortality in patients with HF and AF.^4^

In addition, cognitive decline is related to factors of severity and chronicity of HF and AF, such as NYHA class and pro-BNP level, as well as low left ventricular ejection fraction (LVEF) and physical capacity (PC), also including autonomic imbalance.^1,2,3^ A recent clinical trial confirmed the potential benefits of exercise training for increasing LVEF and PC and improving autonomic imbalance in heart failure patients with reduced ejection fraction and permanent atrial fibrillation (HFAF), which was also suggested in a previous study.^5,6^ The study was a randomized clinical trial that tested whether an exercise training program compared with usual care increased functional capacity, cardiac function, and autonomic response in patients with HFAF.^5^ In that study, we showed that the exercise program is safe, improves quality of life, and promotes clinically important functional changes that are associated with reduced hospitalization rates and can increase survival in these patients.^5^ However, the effect of exercise on other measures of cardiovascular risk factors, such as cognitive function and neurovascular control, is not known among this population.

Regarding exercise type, a recent systematic review and meta-analysis reported the effects of exercise training on cognitive response, with some evidence that suggests positive improvements after exercise.^7^ However, we still lack knowledge about specific populations, for example, HFAF.

This current study details the results of exercise training compared with usual care on the secondary efficacy endpoints of cognitive function, ANSM, FBF, and FVR in patients with HFAF.

## Methods

### Study design and participants

A detailed description of this study design, participants, and intervention was published previously.^5^ Briefly, twenty-six patients with heart failure with reduced ejection fraction and permanent atrial fibrillation were randomly allocated, in a 1:1 ratio, to an exercise training program plus usual care (exercise group) or usual care (control group). The inclusion criteria at screening were age between 50 and 65 years, clinically stable, permanent atrial fibrillation (unsuccessful or indicated for rhythm reversal methods, such as ablation and cardioversion), resting heart rate ≤80 bpm, left ventricle ejection fraction ≤40%, being under optimized clinical treatment and stable during the previous 6 weeks, and able to perform exercise training 3 times a week. Exclusion criteria were patients with pulmonary disease, functional class IV (New York Heart Association [NYHA]), complex ventricular arrhythmia, chronic renal disease, peripheral neuropathy, history of stroke, body mass index (BMI) >30 kg/m^2^, history of smoking, and performing regular physical activity. Participants with a rate >110% of the age-predicted maximum heart rate during CPET were also excluded. The study is registered on ClinicalTrials.gov (NCT03550872). All were approved by the Ethics Committee, conducted following the Declaration of Helsinki, and written informed consent was obtained from all patients.

## Outcomes

### Cognitive Function

Global cognition was assessed using the Brazilian version of the Montreal Cognitive Assessment score (MoCA), which includes an assessment of the cognitive domains of executive and visuospatial, memory, language, and attention.^8^ The MoCA is sensitive for detecting mild cognitive impairment and can be completed in about 10 minutes. The maximum MoCA score is 30 and a score <26 indicates at least mild cognitive impairment. However, graded severity levels of 18–25, 10–17, and <10, are suggested for mild, moderate, and severe cognitive impairment, respectively, although supportive data are lacking.^9^ Therefore, all cognitive impairments will be referred to as mild cognitive impairment. This test has been widely validated and is recommended for HF.^10^ The MoCA was administered by trained research personnel blinded to the protocol.

### Muscle Sympathetic Nerve Activity

MSNA was directly recorded from the peroneal nerve (leg) using microneurography (662C-4 Nerve Traffic Analysis System, The University of Iowa, Iowa City, IA, USA), as previously described.^11^ Muscle sympathetic bursts (bursts per minute) and bursts per 100 heartbeats were identified by visual inspection conducted by a single investigator, blind to the status of study participants (control vs. trained). The normalized average burst amplitude was calculated by assigning the highest absolute burst amplitude recorded at rest as an arbitrary value of 10 and expressing all other burst amplitudes as a percentage of this maximum burst height. Total MSNA activity was then calculated as the product of the mean burst frequency and the normalized mean burst amplitude. Single-unit MSNA was expressed as the total number of peaks per minute (peak frequency) and per 100 heartbeats (peak incidence).^12^

### Forearm Blood Flow

Forearm blood flow (FBF) was measured in the nondominant arm by using strain-gauge venous occlusion plethysmography (EC6 plethysmograph, D.E. Hokanson, Bellevue, WA, USA) as previously described.^11^ Forearm vascular resistance (FVR) was calculated by dividing mean arterial blood pressure by FBF. The reproducibility of FBF measured at different time intervals in the same individual expressed as mL/min/100mL in our laboratory is r. 0.93.

### Exercise Training (ET)

The ET program consisted of 3 weekly morning sessions of 60 minutes of supervised exercise, for 12 weeks, as previously described.^5^ Briefly, the exercise session included 5 min of stretching exercises, 40 min of cycling on an ergometer bicycle, 10 min of strengthening exercises, and 5 min of cool down. The cycling exercise intensity was controlled from 14 to 16 by the Borg Scale.

### Statistical Analysis

Exploratory data analysis and Shapiro-Wilk tests were performed to determine the normality of the data distribution. Results are presented as mean differences with their 2-sided 95%, means and standard deviations, or the median and interquartile range, according to data distribution. Between-group differences at baseline, after the exercise program, and between changes in the continuous variables from before to after intervention were tested with the Student independent *t*-test. Within-group comparisons from before to the end of the intervention were determined with the Student paired *t*-tests. Between-group comparisons for categorical variables were tested with chi-square tests. SPSS v21 software (IBM Inc, Chicago, II) was used to perform these statistical analyses of the results. In all analyses, *P*≤0.05 was considered a significant difference. To get an estimate of the effect size we could expect for the variables in our sample, we relied on results from exercise training studies similar to ours.^9,8,13^ Considering that the results of these studies produced a 6-10% increase in peak VO_2_, we estimated that a total sample size of 13 subjects for each group would be required to provide 85% power to detect a 10% change in peak VO_2_ with two-sides α of 0.05.

## Results

Twenty-six patients completed the protocol study, 13 in the HFAF-trained group and 13 in the HFAF-untrained group. These participants did not report any relevant adverse events during the study. Patients were clinically stable and under medication optimization, at maximum tolerated doses, according to guidelines, and kept the same medication throughout the study. No significant differences in baseline characteristics were observed between the two groups (**Table 1**).

**Table 1.** Baseline characteristics according to the allocated group.

| Parametrs | HFAF-UT (n=13) | HFAF-T (n=13) | *p* value |
| --- | --- | --- | --- |
| Male sex | 13 | 13 | 1,0 |
| Age, years | 58±3 | 58±2 | 0,53 |
| BMI (kg/m<sup>2</sup>) | 26±2,5 | 28±2,7 | 0,06 |
| BSA (m<sup>2</sup>) | 1,9±1 | 2,0±1 | 0,35 |
| LVEF, % | 32±5,5 | 34±4,7 | 0,73 |
| LA, mm | 54±4,7 | 53±7,6 | 0,26 |
| Functional class, NYHA (%) |  |  |  |
| II | 4 | 6 | 0,42 |
| III | 9 | 7 |  |
| Etiology (%) |  |  |  |
| Isquemic | 13 | 13 | 1,0 |
| Medications (%) |  |  |  |
| ACEI ou ARB | 11 | 13 | 0,14 |
| Digoxin | 10 | 8 | 0,40 |
| Espironolactone | 8 | 9 | 0,68 |
| Amiodarone | 1 | 1 | 1,0 |
| Anlodipine | 1 | 2 | 0,58 |
| Beta-blocker | 13 | 13 | 1,0 |
| Diurétic | 13 | 13 | 1,0 |
| Estatíns | 6 | 5 | 0,69 |
| Anticoagulant | 12 | 12 | 1,0 |
| Hypoglycemic | 5 | 6 | 0,69 |
| Exercise |  |  |  |
| Rest Systolic BP, mmHg | 105±11,2 | 105±14,1 | 0,34 |
| Rest Diastolic BP, mmHg | 64±13,1 | 65±13,3 | 0,12 |
| Rest HR, beats/min | 72±6,3 | 75±4,4 | 0,97 |
| Peak HR, beats/min | 148±10,9 | 152±8,2 | 0,41 |
| Peak VO<sub>2</sub> peak, ml.kg.min | 14.2±3.3 | 15.2±2.2 | 0,43 |
| VE/VCO<sub>2</sub> slope | 39±6.3 | 39±4.9 | 0,57 |

HFAF-UT: patients with heart failure and atrial fibrillation untrained; HFAF-T: patients with heart failure and atrial fibrillation trained; BMI: body mass index; BSA: body surface area; ACEI: angiotensin converting enzyme inhibitors; ARB: angiotensin receptor blockers; BP: blood pressure; HR: heart rate; VO<sub>2</sub>: oxygen uptake; VE/VCO<sub>2</sub> slope: relationship between ventilation and carbon dioxide output; LVEF: left-ventricular ejection fraction; LA: left atrium.

At baseline, there were no differences between groups in cognitive function, ANSM, FBF, and FVR. Exercise training, however, significantly improved cognitive function in patients with heart failure and AF (-7.7 [95% CI, -10.1 to -5.4, *P*<0.0001]), but no significant changes were found in the control arm (0.53 [95% CI, -1.3 to 2.4, *P*=0.54]). The change in total MoCA score was significantly different between groups at 5.4 (95% CI, 1.8 to 9.0, *P*=0.004) (**Figure 1**).

**Figure 1.**
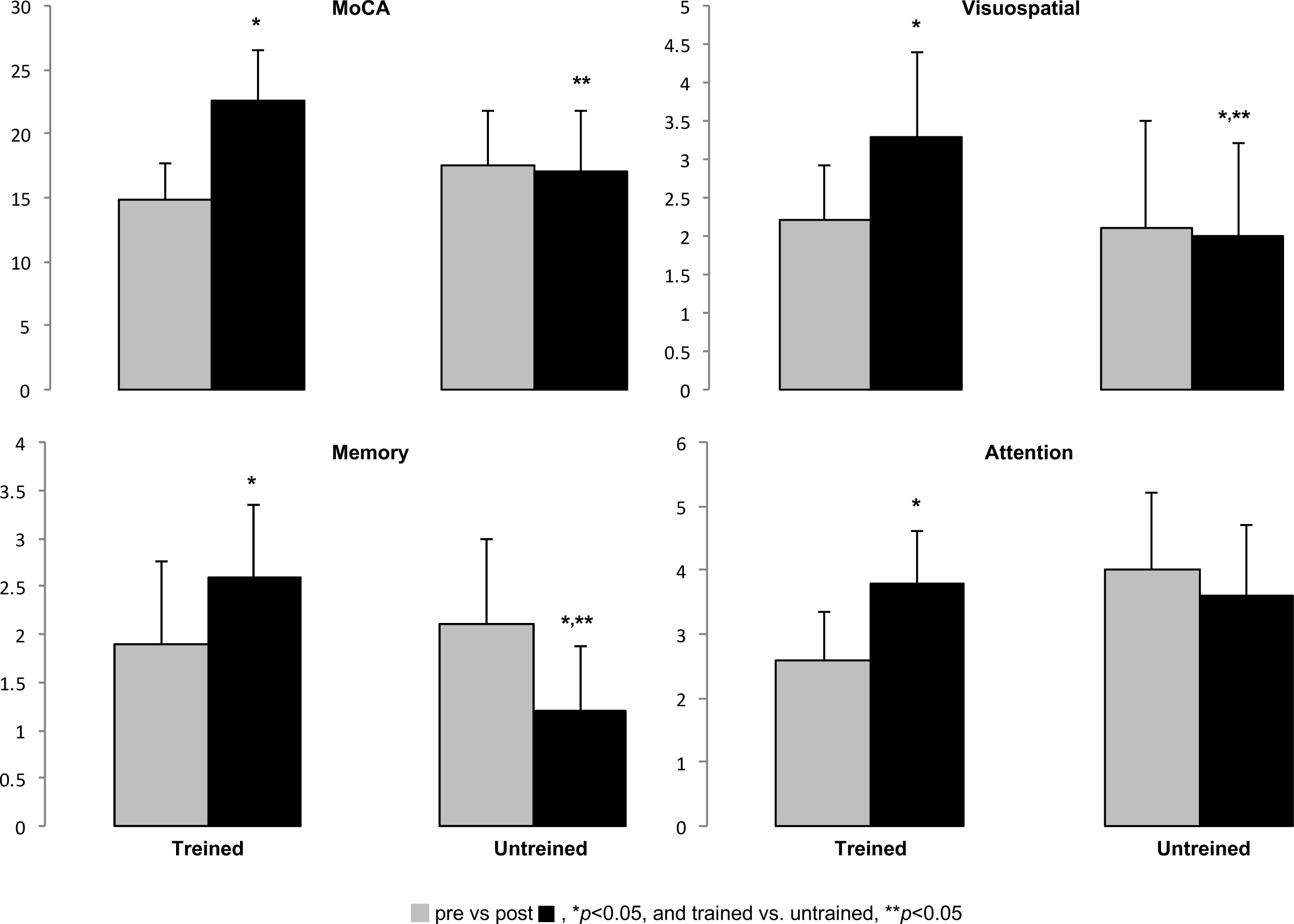
Montreal Cognitive Assessment score (MoCA), visuospatial, memory, and attention function in trained and untrained HFAF. Exercise training significantly improves cognitive function. Significant difference pre vs. post, \**P*<0.05, and trained vs. untrained, \*\**P*<0.05.

Patients in the exercise arm significantly decreased MSNA by 17.6 (95% CI, 12.6 to 22.7, *P*<0.0001), whereas patients in the control arm significantly increased MSNA by -6.6 (CI 95%, -10.8 to -2.5, *P*=0.004). This behavior resulted in a significant difference between groups of -25.6 (95% CI, -32.3 to -18.8, *P*<0.0001) (**Figure 2**).

**Figure 2.**
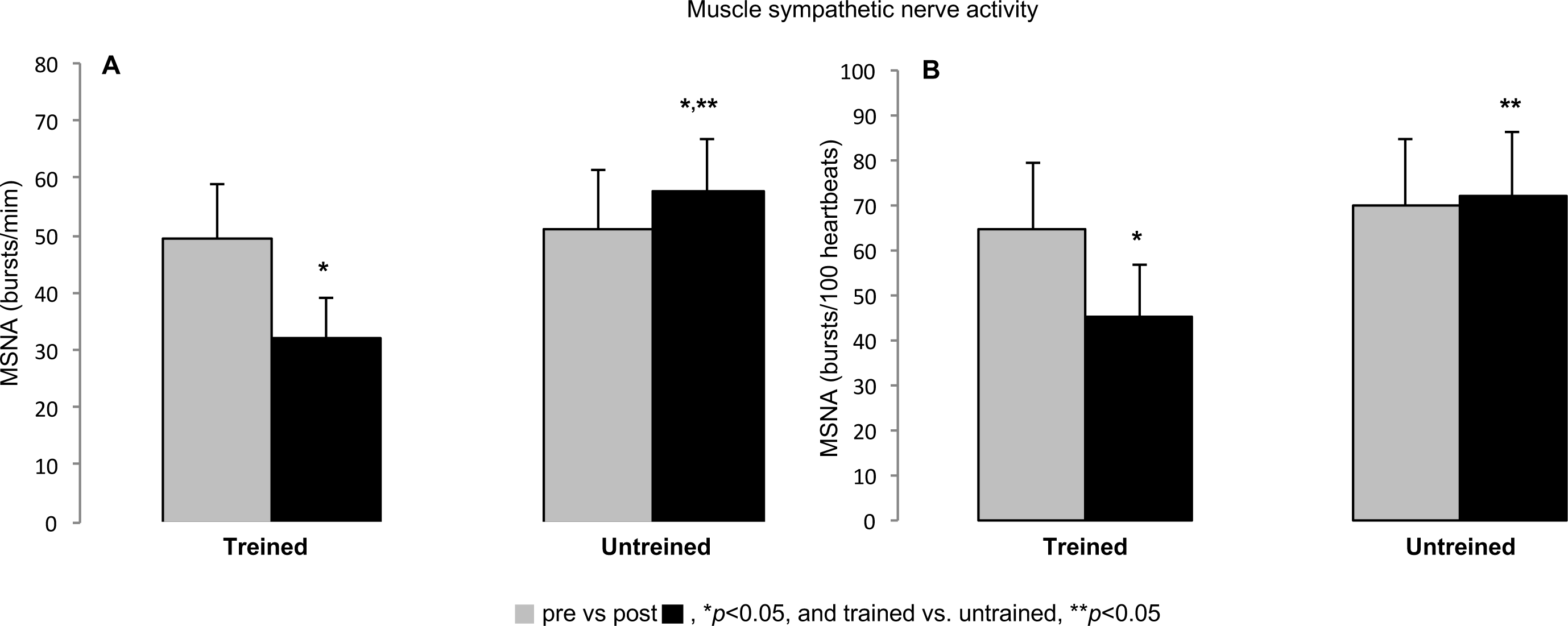
Muscle sympathetic nerve activity (MSNA) was quantified as bursts/min (A) and bursts/100 heartbeats (B) in trained and untrained HFAF. Exercise training significantly reduced MSNA levels. However, MSNA levels increased significantly in the untrained group. Significant difference pre vs. post, \**P*<0.05, and trained vs. untrained, \*\**P*<0.05.

The change in FBF was significantly different in the exercise arm at -1.1 (95% CI, -1.4 to -0.8, *P*<0.0001), whereas no change in the control arm was found for FBF (0.1 [CI 95%, -0.7 to 0.4, *P*=0.14]. In addition, there was a significant difference between groups of 1.1 (CI 95%, 0.6 to 1.6, *P*<0.0001) (**Figure 3A**).

**Figure 3.**
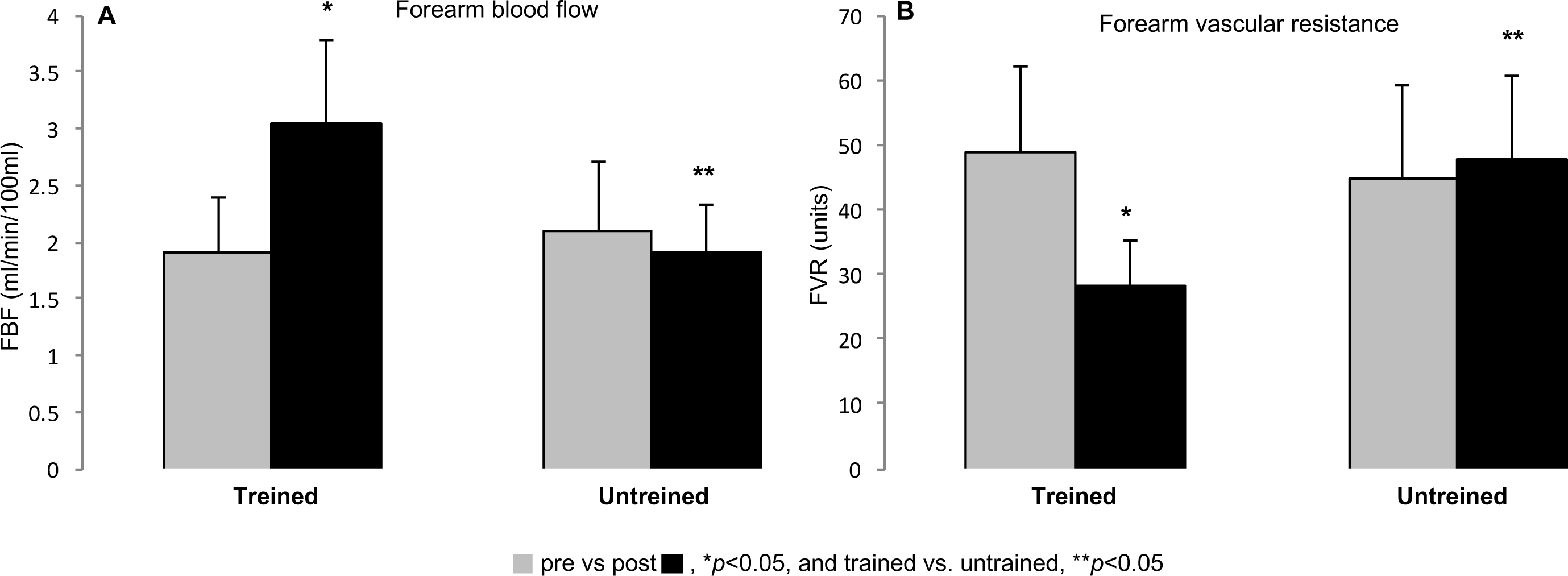
Forearm blood flow (FBF) (A) and forearm vascular resistance (FVR) (B) in trained and untrained HFAF. Exercise training significantly increased FBF, and significantly decreased FVR. FBF and FVR were not significantly changed in the untrained group. Significant difference pre vs. post, \**P*<0.05, and trained vs. untrained, \*\**P*<0.05.

Exercise training significantly reduced the FVR by 20.7 (95% CI, 16.1 to 25.9, *P*<0.0001). No significant change was observed in untrained patients (-3.1 [95% CI, - 9.4 to 3.0, *P*=0.29]. The decrease in the exercise group resulted in a significant difference between groups of -19.6 (95% CI, -28.0 to -11.1, *P*<0.0001) (**Figure 3B**).

## Discussion

The previous clinical trial showed that an exercise training program resulted in improvements in cardiac function, functional capacity, and quality of life, and reduced resting and recovery heart rate in HF with reduced ejection fraction and permanent AF.^5^ Although the effects of exercise interventions are increasingly recognized in the treatment of HF and AF,^5,6,11^ to our knowledge, this is the first study to document the beneficial effects of an exercise program on cognitive function and neurovascular control among patients with HFAF. These results, together with the primary findings, are clinically promising because they are associated with reduced cardiovascular risk factors and hospitalization, as well as improved physical well-being and the achievement of mental health benefits in this population.

The presence of AF in patients with HF was associated with worse cognitive performance compared with patients with HF without AF.^1,2,3,4^ One possible mechanism is that loss of atrioventricular synchrony leads to cerebral hypoperfusion, suggesting that AF exacerbates cognitive dysfunction in patients with HF, possibly through a greater reduction in cerebral blood flow. Moreover, cerebral microbleeds may play a role in cognitive decline, especially in patients with HFAF who are being treated with anticoagulation.^1,2^ However, older AF patients who are on anticoagulant therapy and have no cognitive impairment are at a lower risk of cerebral microbleeds compared with those who do have cognitive decline.^13^ Another important fact is that, in some specific cases, the use of direct oral anticoagulant therapy in patients with HFAF may negatively impact cognitive performance.^14,15^

On the other hand, physical activity in adults is known to play an important role in the protection against cognitive decline and dementia through processes of neuroplasticity.^16,17^ Our results demonstrate that exercise training increases FBF and decreases FVR. This improvement in vascular conductance may result from increased endothelial function due to the acceleration of blood flow during physical exercise, causing the release of brain-derived neuroplastic factors.^18,19^

The brain is involved in the progression of HF through sympathetic activation and regulation of fluid homeostasis, which can affect cerebral autoregulation and decrease cognitive function.^20^ In patients with irregular heartbeats, such as atrial fibrillation, sympathetic nerve activity may worsen the severity of heart failure and lead to cognitive decline.^1,2^ Studies have demonstrated a link between MSNA and cognitive unction in patients without dementia and have shown that lower cognitive performance is associated with significantly greater cardiac sympathetic function and lower parasympathetic function.^21^

In the previous study, we showed that exercise training significantly reduces MSNA and increases muscle blood flow in patients with HF.^6,11^ The present study confirms these findings and provides the knowledge that reducing MSNA and vasoconstriction after exercise training may be associated with improved cognitive function in patients with HFAF. The mechanisms by which exercise training reduces sympathetic nerve activity may be, in part, due to the restoration of baroreflex control, the decrease in angiotensin II in the central nervous system, and the improvement of chemoreflex control due to the increase in nitric oxide synthesis.^6,11,21^ These results support the concept that exercise training is an important modality to improve cognitive function in HFAF-trained, which was not found in the untrained group.

Cognitive decline and low physical fitness are critical issues among older adults; however, older adults with greater physical fitness performed as well as younger adults on cognitive function tests.^22,23^ Exercise training in the present study shows beneficial effects on cognitive function. We believe that our study is the first to report the association between exercise training and MoCA score in HF with reduced ejection fraction and permanent AF. Cognitive impairment is defined as an MoCA score <26; on the other hand, there is a MoCA cutoff score ≥23 that can predict a return to normalized cognitive function with exercise training among elderly outpatients.^24^ At baseline, cognitive impairment was present in the HFAF patient sample (mean MoCA score 17.56). After 12 weeks of exercise training, only the HFAF-trained group showed improvement in their MoCA score, which increased to 22.62. The specific mechanisms underlying the improvement in cognitive function after exercise training in HFAF with cognitive decline are still being explored.^1,2,3,9,10^ However, the improvement in cognitive function after exercise training may be related to the set of benefits demonstrated in this study together with the results of the previous study to increase brain-derived neuroplastic factors.

Furthermore, several factors, including biological, psychological, cultural, and environmental factors (such as aging, lack of physical activity, chronic illnesses, and low levels of education) can have a negative impact on cognitive ability.^22^ Therefore, exercise training may be a useful addition or alternative method to improve cognitive function in patients with HFAF.

## Clinical Implications

Research shows a connection between physical performance and cognitive function in healthy people. Each 1 MET increase in physical capacity is linked to a 7% reduction in the risk of cognitive impairment.^25^ Our study supports this finding, highlighting the importance of regular exercise training for patients with HFAF. In addition, improved cognitive function can increase patient self-care and adherence to treatment, resulting in lower hospitalization and mortality rates in HF patients with reduced ejection fraction and permanent AF.^4^ However, further research should prioritize investigating the underlying pathogenesis and mechanisms to gain a better understanding of the cognitive dysfunction experienced by patients with heart failure and atrial fibrillation. Although previous studies have been conducted, the precise pathophysiology of cognitive impairment in this patient group is yet to be fully understood.

## Limitations

Although this study provides valuable insights, it is important to note its limitations. Firstly, the MoCA has varying thresholds for screening cognitive impairment in patients with HF and cardiovascular disease.^26^ A score below 26 may over-pathologize cognitive impairment in these populations.^16^ Secondly, the MoCA relies heavily on abilities learned in school, and low scores may indicate educational limitations or pathology.^1,2^ However, we did not evaluate education levels in this sample. Third, we did not perform cerebral computed tomography scans or other imaging techniques that could have provided relevant information, particularly concerning vascular cognitive impairment. Fourth, of the 196 people evaluated, only 50 were women, as previously described.^5^ Unfortunately, 18 of them did not meet the inclusion criteria, 10 of them had a BMI > 30 kg/m^2^ and 12 refused to participate for personal reasons. Therefore, these results cannot be applied to women with HFAF. Finally, recent evidence shows that multi-component exercise training, which includes motor and physical components, may enhance cognitive performance better than traditional exercise.^26,27^ Unfortunately, our exercise training program did not include these components, such as balance, mobility, coordination, agility, proprioception, and reaction time.

Despite these limitations, this is the first study that evaluated the influence of exercise training on cognitive and neurovascular performance in HF with reduced ejection fraction and permanent AF.

## Conclusion

The current study aimed to investigate the potential significance of exercise training for cognitive and neurovascular function in patients with HFAF. The main finding is that exercise training can have beneficial effects on cognitive and neurovascular function in patients with HFAF. The associations with better neurocognitive performance may indicate a potential protective effect of exercise training, but more research is needed to confirm this. Until further studies are conducted, patients with HFAF can be encouraged to practice exercise training.

## Author contribution

GVG: study concept and design, analysis and interpretation of data, and preparation of manuscript. PRC: study concept and design, and preparation of manuscript. PRSS: analysis and interpretation of data, and preparation of manuscript. RVGM: acquisition of subjects and/or data. LSA: study concept and design, acquisition of subjects and/or data. EAB: study concept and design and interpretation of data. All authors gave final approval and agreed to all aspects of the work ensuring integrity and accuracy. The current study is presented honestly without fabrication, falsification, or inappropriate data manipulation. The authors declare no potential conflicts of interest concerning the research, authorship, and/or publication of this article.

## Data Availability

The data presented in this study are available on request from the 415 corresponding author.

